# Sex-Dependent Relationships Between PFAS and Placental Transcriptomics Identified by Weighted Gene Co-Expression Analysis

**DOI:** 10.1101/2025.06.23.25330157

**Authors:** Cynthia Perez, Kyle Campbell, Dana Boyd Barr, Kartik Shankar, Clark Sims, Kevin J. Pearson, Aline Andres, Todd M. Everson

## Abstract

**Background:** Per– and polyfluoroalkyl substances (PFAS) are environmental toxicants associated with adverse neonatal outcomes. The exact mechanisms by which PFAS impairs neonatal health are undefined, but the placenta is a likely target.

**Objective:** We applied a systems biology approach to identify placental RNA co-expression modules (gene sets) associated with PFAS exposure and birth weight.

**Methods:** Placental tissue samples (n = 147) from the GLOWING study underwent RNA-sequencing, and PFAS concentrations were quantified using liquid chromatography-tandem mass spectrometry. We constructed a weighted gene co-expression network using Spearman correlations across 15,028 transcripts, identifying 20 gene modules. Linear regression models were used to examine associations between PFAS and module eigengenes, adjusting for potential confounders. Effect modification by fetal sex was also tested.

**Results:** One module showed a negative association with perfluorononanoic acid (PFNA; β = – 0.012, q = 0.009). This association was sex-specific, with the sexes exhibiting varied PFAS associations but similar directional effects. Genes within the PFNA-associated module were involved in histone modification (q ≤ 0.05) and were enriched for targets of the Vitamin D Receptor (VDR), a transcription factor previously linked to PFAS.

**Discussion:** Our research indicates that prenatal exposure to PFNA influences placental gene expression differently based on sex, which may affect insulin growth factor signaling and histone modification. The presence of VDR in this module and the transcription enrichment analysis align with previous findings regarding PFAS and VDR interactions. This module related to PFNA could shed light on the molecular pathways connecting PFAS exposure to health outcomes in neonates.

## Introduction

Per– and polyfluoroalkyl substances (PFAS) are characterized by a repetitive chain of carbon and fluorine bonds, along with a distinctive functional group.^1^ This structural composition renders PFAS a practical choice for repelling water and grease from a wide range of products. This same structure also contributes to its extended half-life and persistent environmental contamination.^2, 3^ Heightened exposure to PFAS can present health risks such as thyroid dysregulation, neurological impairment, and increased susceptibility to cancer.^4–8^ In an effort to mitigate these risks, the U.S. Environmental Protection Agency reached an agreement with eight U.S. companies to cease the use and production of PFAS with the lengthiest history of use, specifically perfluorooctanoic acid (PFOA) and perfluorooctanesulfonic acid (PFOS).^9^ However, the agreement did not address the replacement PFAS the companies would use nor did it address remediation of the present environmental contamination of PFOA and PFOS. PFAS are a critical environmental health concern, with exposure being especially concerning during early human development.

Our group and others have shown that PFAS accumulate in placental tissue during pregnancy, and that prenatal exposure to PFAS is associated with molecular responses, low birth weight, impaired cardiovascular health, and increased dyslipidemia in neonates.^10–12^ Among the molecular mechanisms of growing interest are peroxisome proliferator-activated receptors (PPARs), a nuclear receptor superfamily crucial for embryo implantation and development of the placenta.^13^ PPARs can regulate the expression of genes essential for cell differentiation and various metabolic processes like glucose homeostasis.^13^ Research suggests all three PPAR isomers are affected by PFAS, with PPARα being a particularly common target.^14^ Other nuclear receptors, including androstane receptor and pregnane X, are also impacted by PFAS.^15, 16^ The relationship between PFAS and the activity of key transcription factors, like nuclear receptors, may influence the expression of a plethora of genes. Therefore, it is advantageous to utilize a systems biology approach to better understand impacts of PFAS on transcriptional dysregulation.

Weighted gene co-expression network analysis can model complex interactions between expressed genes and discover emergent system properties.^17^ These networks utilize global gene expression measurements (RNA sequencing or microarray-based data) to reveal sets of genes which may share the same transcriptional regulatory system, members of the same biological pathways, or that are functionally related.^18^ Weighted gene co-expression networks may be useful in characterizing system-wide placental biological activity and/or disruptions caused by PFAS, providing novel insights into how PFAS affect placental function.

Our previous research indicated that placental PFAS influenced DNA methylation, accelerated gestational age in females, and correlated with a reduced predicted syncytiotrophoblast proportion in males. Consequently, we aimed to uncover functional gene networks impacted by PFAS.^19, 20^ We applied Weighted Gene Correlation Network Analysis (WGCNA) to identify PFAS-associated co-expressed gene sets (or modules) from placental RNA sequencing profiles. Emerging research, including our own, suggests that in utero exposure to PFAS may lead to sex specific developmental effects in vivo.^19, 21, 22^ Therefore, we hypothesized that PFAS-associated changes in gene expression may be sexually dimorphic. Due to regions of high sequence similarity between the X and Y chromosomes, we employed a sex complement alignment method to identify true sex differences.^23^ We found one module composed of mainly sex-linked genes to be associated with perfluorononanoic acid (PFNA). Most PFAS do not appear to affect system-wide gene expression in the placenta, but PFNA does only in a sex specific manner.

## Methods

### Study Population

The Glowing Cohort was established as part of the VICTER consortium project (R01 ES032176) to investigate the effects of in utero PFAS exposure on placental omics. Between 2010 and 2014, 300 pregnant participants were recruited from the Little Rock, Arkansas, metropolitan area. Further details on recruitment criteria can be found in prior work.^19, 20^ In short, the cohort consists exclusively of non-smoking participants over the age of 21 without complicated births or serious medical conditions. At enrollment, participants were categorized as lean or overweight based on pre-pregnancy BMI, which was determined by self-reported height and weight.

### Placenta collection and quantification of PFAS

A subsample of 150 participants provided sufficient placental tissue for RNA sequencing and PFAS quantification. Placental physical properties (e.g., weight) were measured within two hours of collection, after the removal of the umbilical cord and fetal membranes. Approximately 1 gram of placental tissue from six random sites within the villous core was pulverized in liquid nitrogen and flash frozen. We have described the quantification of the 17 PFAS chemicals in our population sample via liquid chromatography-tandem mass spectrometry (LC-MS/MS) elsewhere.^19, 20^ We restricted our analysis to the 5 most well-detected PFAS with above limit of detection rates greater than 65% among all samples: perfluorooctane sulfonate (PFOS), perfluorohexane sulfonic acid (PFHxS), perfluorononanoic acid (PFNA), Perfluorooctanoic acid (PFOA), and perfluorodecanoic acid (PFDA). Samples were randomized across runs to reduce potential batch effect. The limit of quantification (LOQ) values for PFAS are represented in **Table 1** along with concentration ranges. Values less than the LOQ were replaced with 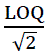 to minimize potential bias.^24^

**Table 1:** Summary statistics of 17 PFAS chemicals measured in N = 147 of which RNA-sequencing profiles and had sufficient yield for PFAS quantification.

| PFAS | Geometric Mean | Geometric SD | LOQ Range | >LOQ (%) |
| --- | --- | --- | --- | --- |
| PFOS | 0.375 | 2.010 | 0.021 - 2.038 | 99% |
| PFNA | 0.049 | 1.866 | 0.007 - 0.222 | 98% |
| PFHxS | 0.047 | 2.795 | 0.007 - 0.816 | 83% |
| PFOA | 0.051 | 2.890 | 0.012 - 0.416 | 72% |
| PFDA | 0.025 | 2.461 | 0.007 - 0.108 | 71% |
| PFUnDA | 0.020 | 2.168 | 0.013 - 0.138 | 31% |
| PFHpA | 0.007 | 1.816 | 0.006 - 0.184 | 20% |
| HFPO-DA | 0.015 | 1.495 | 0.014 - 0.420 | 4% |
| PFHxA | 0.007 | 1.241 | 0.007 - 0.237 | 2% |
| MePFOSAA | 0.073 | 1.240 | 0.071 - 0.347 | 2% |
| PFBS | 0.007 | 1.151 | 0.007 - 0.039 | 1% |
| PFDoDA | 0.009 | 1.143 | 0.009 - 0.032 | 1% |
| PFOSA | 0.071 | 1.061 | 0.071 - 0.145 | 1% |
| PFPeA | 0.014 | 1.000 | 0.014 - 0.014 | 0% |
| PFHpS | 0.014 | 1.000 | 0.141 - 0.141 | 0% |
| PFDS | 0.035 | 1.000 | 0.035 - 0.035 | 0% |
| EtPFOSAA | 0.071 | 1.000 | 0.071 - 0.071 | 0% |

### RNA Sequencing Data Process

To conduct transcriptomic analyses, RNA was isolated from pulverized heterogenous placenta samples. Total RNA was extracted from the placental samples using TRI reagent, followed by purification using RNeasy mini columns and deoxyribonuclease digestion. Directional RNA-sequencing libraries were created using NebNext Ultra reagents and sequenced on an Illumina HiSeq 4000 platform, generating approximately 30 million reads per sample. Before and after trimming with Trim Galore, FASTQC and MultiQC were respectively used to assess individual and aggregated sample quality.^25–27^ Three samples were excluded because they lacked the yield for both PFAS quantification and RNA sequencing. The trimmed sequences were then aligned to the Gencode GRCh38.p12/hg38 gene annotation using STAR in a sex complement manner where the Y pseudoautosomal regions (PAR 1 and PAR 2) are ignored for males and the Y chromosome is completely ignored for females.^23, 28^ RNA sequencing alignment files were then sorted and indexed using bamtools.^29^ Mapped reads were quantified and summarized into counts using featureCounts.^30^ QC metrics required that samples have more than 12.5M and fewer than 90M sequences after trimming, and less than 30% of reads that deviate from the normal distribution of per-sequence GC content based on the FastQC report.^31^ All 150 samples passed QC metrics. The analytic dataset consisted of 15,028 RNA transcripts, including 513 X-linked genes and 16 Y-linked genes. Additionally, to address potential confounding by batch variation effects, we corrected gene expression counts with the ComBat function from the sva package (v 3.52.0), while protecting the covariates in our primary models (maternal age, maternal education, gestational age at delivery, gestational weight gain, biological sex of the neonate, maternal BMI category at enrollment, and the first principal component of RNA-seq data).^32^

### Statistical Analyses

#### Weighted Gene Co-expression Network Analysis

All analyses were performed using R (version 4.2.2). To identify system-wide gene expression patterns, we used Weighted Gene Co-expression Network Analysis (WGCNA), which aims to discern clusters of genes that display comparable patterns of expression, indicating potential functional relationships among them in the WGCNA R package (v 1.73).^17^ To prepare gene expression data for WGCNA, we normalized expression count data with upper quartile normalization and variance-stabilizing transformation using the R package TRAPR.^33^ These normalization methods were chosen to optimize co-expression module identification via WGCNA.^34^ We chose to employ a signed network approach, which focuses on the strength of positive gene-gene correlations to draw network connections. While there are other network types, signed networks provide ease of biological interpretability as compared to networks that include negative correlations.^35^

To construct the signed network, we utilized the upper-quartile normalized, variance-stabilized, and batch-corrected mRNA-seq data to calculate pairwise gene-gene Spearman correlations across 15,028 genes and 147 participants. These correlation values were applied in the following equation to ascertain the similarity, *s*, between genes *i* and *j*: s_ij_ = 0.5 + 0.5 ∗ cor(i, j), which quantifies the relationship between genes and penalizes negative correlations. Consequently, negative correlations had fewer connections in the adjacency matrix, a_ij_ = |s_ij_|^β^, where β is a parameter determining the scale-free topology of the network. The adjacency matrix outlines the network’s structure before the clustering process, aimed at establishing co-expression patterns among genes.

To achieve a scale-free topology, we utilized Spearman correlation and selected β = 5 based on the pickSoftThreshold function. We calculated eigengenes, first principal component of each module, to represent overall module expression. The package arbitrarily assigned colors to each identified module. Grey was assigned to genes that did not form a distinct cluster.

#### Differential Expression Analysis

To address the right skewed distributions of PFAS measures, PFAS levels were natural log transformed. All models included the following covariates: maternal age (continuous), maternal education (categorical), gestational age at delivery (continuous), gestational weight gain (continuous), biological sex of the neonate (dichotomous), maternal BMI category at enrollment (dichotomous), and the first principal component of RNA-seq data (continuous). These have been previously documented as confounders in studies focused on in utero PFAS exposure.^12, 36^ The first principal component was included in our models to account for RNA-sequencing technical differences or other unmeasured sources of variation (**Supplemental Figure 1**).

In addition to WGCNA analyses, we conducted differential expression analyses by empirical Bayes-based models through the limma R package (v 3.60.6) to determine if individual gene expression was associated with PFAS concentration. We assessed the effects of PFAS on all 15,028 genes and additionally honed our analysis to the 519 gonosomal transcripts in sex-stratified models where we dropped Y-linked genes for female specific analysis. P-values were adjusted using Benjamin-Hochberg correction (15,028 tests per PFAS).

#### Module Eigengenes and PFAS Analysis

We investigated potential associations between placental PFAS concentrations and changes in the module eigengenes using multiple linear regression. For these models, PFAS concentrations served as the independent variables and the module eigengenes were treated as the dependent variables, while controlling for confounders. We also explored whether there were significant interactions between PFAS and sex on module eigengene levels by including a sex by PFAS cross-product term in our models. To investigate potential effect modification by sex in the association between PFAS and module eigengenes, we then stratified the models by sex. Lastly, we explored the associations between module eigengenes and birth outcomes (gestational age, birth weight, and birth length) to examine whether the same modules impacted by PFAS also associate with these characteristics.

Gene set enrichment analysis was performed for each module using the R package gprofiler2 with the database Gene Ontology (GO).^37–39^ The REVIGO tool was used to filter out redundant terms and to visualize enrichment.^40^ To identify possible reasons for co-expression within each module, we utilized a placental transcription factor (TF) network to determine enriched TFs through a Fisher’s exact test.^41^

## Results

### Sample description

In our study, 150 placental RNA-sequencing samples passed initial QC. We excluded 2 samples with insufficient yield for PFAS quantification and 1 sample that did not cluster with the reported sex assigned at birth, resulting in a final sample size of N=147. The demographic characteristics of the study population are summarized in **Table 2**. Maternal ages ranged from 22 to 42 years old, with a mean age of 30.59 years at the time of birth. The sex distribution of neonates had a greater proportion of male newborns (62.59%). Participants primarily self-reported their ethnicity and race as non-Hispanic White (80%).

**Table 2:** Demographic characteristics of N=147 participants.

| **Maternal and Infant Characteristics** | **Mean (SD)** |
| --- | --- |
| Gestational Weight Gain (kg) | 11.86 (4.22) |
| Gestational Age (weeks) | 39.33 (0.84) |
| Maternal Age (years) | 30.52 (3.46) |
| Birth Weight (kg) | 3.54 (0.43) |
| Birth Length (cm) | 51.28 (2.31) |
|  | **N (%)** |
| BMI Enrollment Category |  |
| Mother with normal weight | 73 (50) |
| Mother with overweight or obesity | 74 (50) |
| Sex of Infant |  |
| Female | 60 (41) |
| Male | 87 (59) |
| Maternal Education |  |
| College Graduate | 56 (38) |
| Graduate Training or Degree | 44 (30) |
| High School Graduate, GED, Associate, Partial College, or Specialized Training | 47 (32) |
| Race of Infant |  |
| White | 121 (82) |
| Non-White | 26 (18) |
| Ethnicity of Infant |  |
| Hispanic | 5 (3) |
| Non-Hispanic | 142 (97) |

Among the 17 analyzed PFAS compounds, PFOA, PFHxS, PFNA, PFDA, and PFOS were detectable in more than 65% of samples and were the focus of this analysis. Over 50% of samples exceeded the LOQ for all five chemicals, and only one sample had concentrations lower than the LOQ for all five PFAS. Concentrations of these five compounds were moderately correlated (p-values < 0.05) with each other, with Spearman correlations ranging from 0.3 to 0.71 (**Supplemental Figure 2**). The subsample used in this analysis produced similar demographic and PFAS concentration metrics comparable to those in our previous publications.^19, 20^

### Differential Expression

We first performed differential gene expression analysis to identify individual genes whose expression is associated with any of the 5 well-detected PFAS. This was achieved through a series of empirical Bayes-based models, where the dependent variable was the expression of one gene, and the independent variable was a PFAS congener and adjusted for covariates. We found no association between PFAS and gene expression after Benjamin-Hochberg correction. However, we hypothesized that there may be sex-specific associations among the sex-linked genes. In the female stratum, seven genes were significantly associated with a PFAS chemical (adjusted for 513 tests). Genes *DDX3X* (q-value = 0.035), *KDM6A* (q-value = 0.015), and *XIST* (q-value =0.005) were positively associated with PFNA while *VAMP7* (q-value = 0.001) and *CD99* (q-value = 0.041) were negatively associated with PFNA (**Figure 1A**). The expression of genes *BCLAF3* (q-value = 0.05) and *PRKX* (q-value = 0.021) were positively associated with PFOS (**Figure 1B**). In the male stratum, no differential expression was observed.

**Figure 1:**
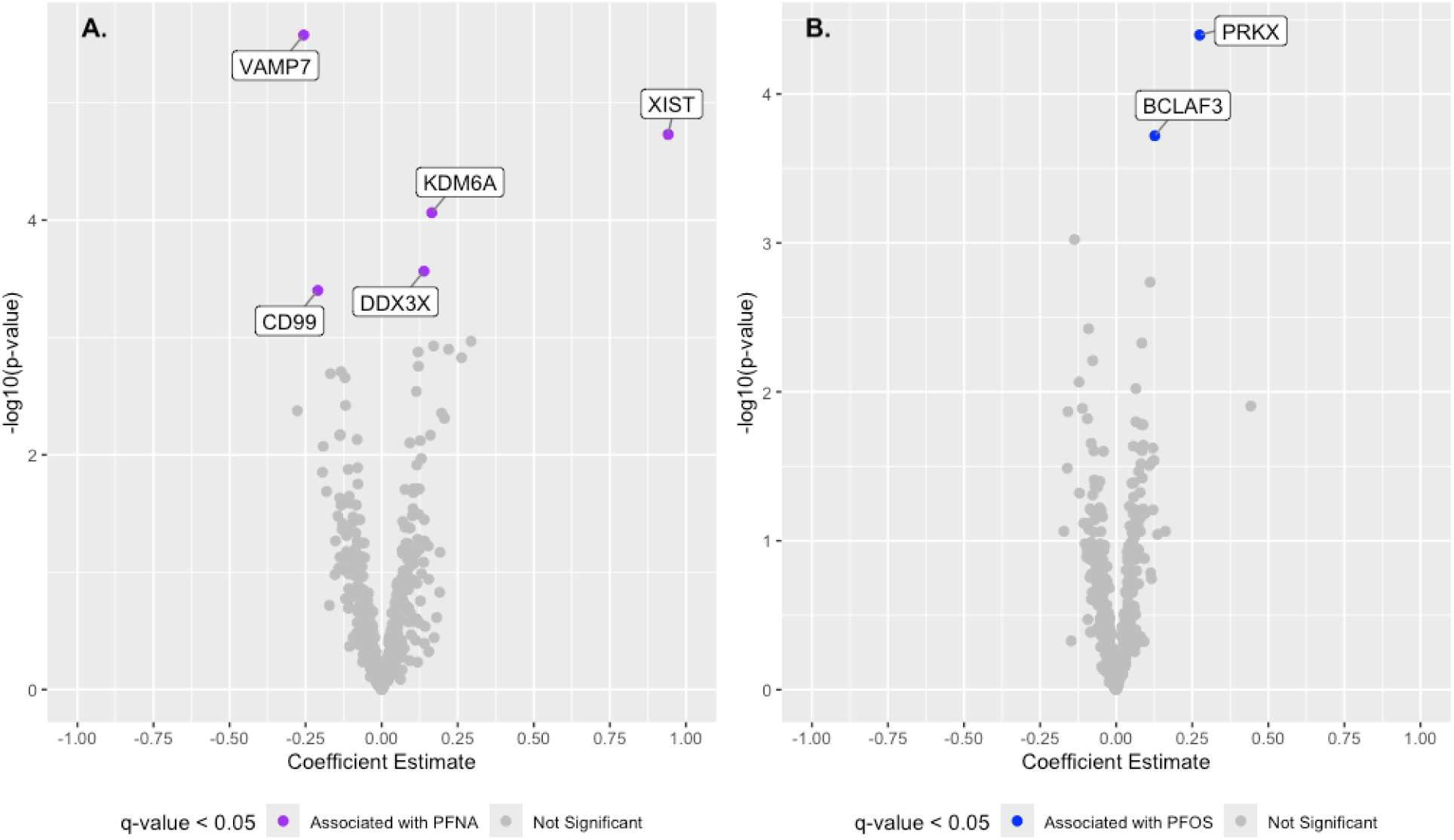
Differential expression analysis in the female strata conducted between 513 X-linked mRNA transcripts, for PFNA. (A) and PFOS (B). The x axis is the coefficient estimate in response to PFNA or PFOS. The y-axis is the log_10_ transformed p-value. Significant differentially expressed genes (DEGs) are denoted in purple for PFNA or blue for PFOS.

### Placental Gene Co-Expression Network

Next, we aimed to identify placental gene expression modules and their associated biological functions in our dataset, which could be impacted by PFAS exposure. For this, we used weighted gene co-expression network analysis (WGCNA), which identified 20 co-expressed modules, and excluded grey which contained 214 genes that were not members of a co-expressed module. The number of genes within each module ranged from 37 to 8328. The proportion of variance explained by the eigengenes, the first principal component of each module, ranged from 31.1% (brown) to 94.2% (light green) with an average proportion of 53.9%.

Gene set enrichment analysis was performed on each module separately, using the GO consortium (**Supplemental File 1**).^37^ Each module exhibited enrichment for at least one GO term within one of the three biological domains: molecular function, biological process, or cellular compartment. Module-specific enrichment yielded multiple redundant terms that were then summarized and represented as tree maps by REVIGO. For simplicity, we labeled each module with the most pertinent biological process (**Figure 2**).^40^

**Figure 2:**
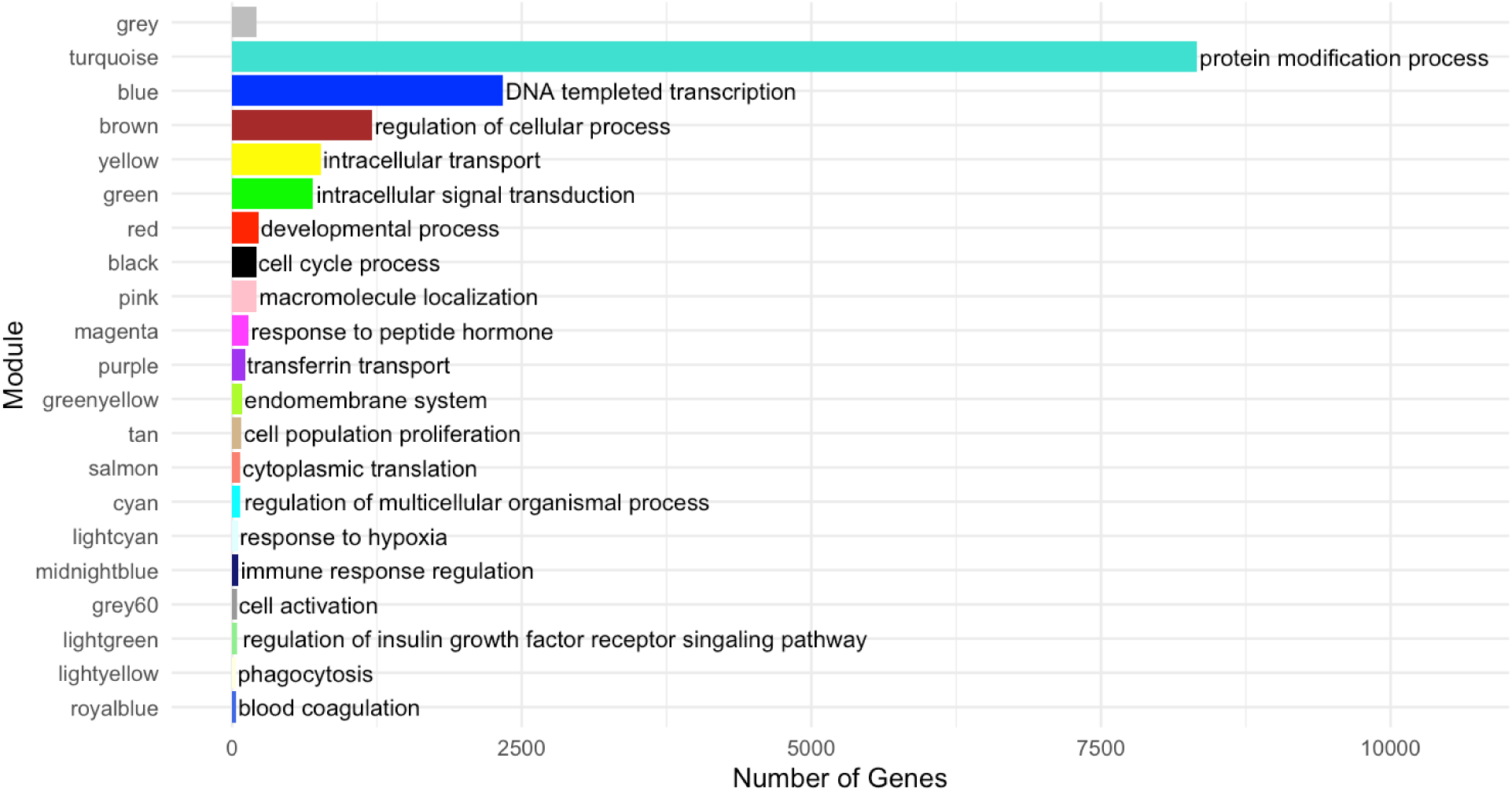
Weighted gene co-expression network analysis identified 20 co-expressed modules, and each was assigned a color (y-axis). The number of genes in each of the modules (x-axis) ranged from 37 to 8328 genes. We labeled each module with the most pertinent biological processes provided by REVIGO. Grey includes 214 genes not co-expressed.

To explore the reasons for co-expression within the modules, we conducted a transcription factor (TF) enrichment analysis using a TF network specific to the placenta.^41^ TFs that targeted genes within a module were assigned an enrichment p-value and q-value for each TF (**Supplemental File 2)**. For modules blue and green, the enriched TFs were not significant after adjustment. The magenta module had the highest number of enriched TFs (q-value < 0.05) with 39, while the light green module had the fewest, with only 2.

We also assessed whether any module eigengenes were related to birth outcomes. Eight modules were associated with at least one of gestational age, birth weight, or birth length (Pearson correlation, nominal p-value < 0.05) (**Supplemental Figure 3**). However, the correlation values between the eigengenes and the birth outcomes were generally weak (r range: –0.2 – 0.19) and these associations were not significant after correction for multiple testing. When these relationships were modeled with linear regression and confounding variables were included, we found no evidence of an association between module eigengenes and the birth outcomes that we tested.

### Module and PFAS associations

We individually modeled the relationship between each of the 20 co-expressed module eigengenes and each of the 5 PFAS, adjusted for confounders. After false discovery rate adjustment, one eigengene, light green, was significantly associated with PFNA (β = –0.012, q-value = 0.009). This module contains 40 genes, of which 33 are sex-linked (17 X-linked and 16 Y-linked). Chromosome 20 is the only autosome with more than one gene represented within the light green module (**Table 3**). Interestingly, consistent with our sex-specific differential expression analysis, the light green genes *KDM6A*, *PRKX*, and *XIST* were also significantly differentially expressed among females.

**Table 3:**
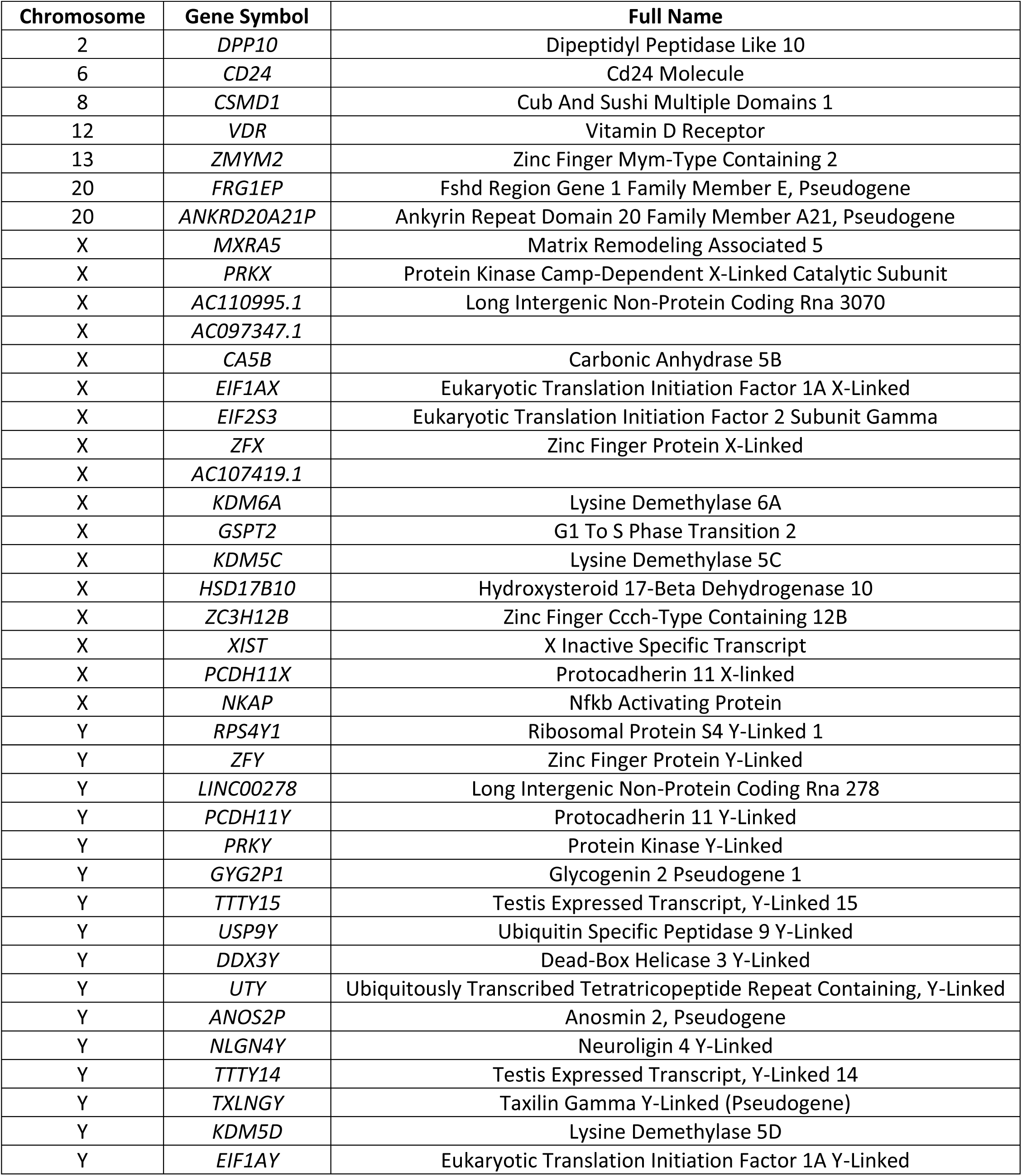
The 40 light green genes featuring their chromosome location. The module includes 33 sex linked genes (17 X-linked and 16 Y-linked). Gene annotation file included long coding RNAs with no official name as identified by the HUGO gene name nomenclature committee.

#### Investigating Sexual Dimorphism of PFAS Associations

Given that many genes in light green were located on sex chromosomes, and PFAS are hypothesized to have sex-specific effects, we investigated whether there were statistical interactions in linear models between sex and PFAS exposure on each of the eigengenes. We identified a significant interaction between PFNA and sex for the light green eigengene (β = 0.023, q-value = 0.01, **Table 4**). Several other PFAS exhibited nominally significant (raw p < 0.05) interactions for light green.

**Table 4:** Linear regression analyses examining PFAS as the independent variable and the eigengene of the light green module as the dependent variable (adjusted for confounders), both with and without an interaction term between a PFAS chemical and the biological sex of neonate.

| Main Effect |  |  |  |  |
| --- | --- | --- | --- | --- |
| PFAS | 95% CI | Estimate | p-value | q-value |
| PFOA | [-0.005,0.002] | -0.001 | 0.465 | 0.952 |
| PFHxS | [-0.004, 0.004] | -0.0002 | 0.921 | 0.994 |
| PFNA | [-0.017, -0.006] | -0.012 | <0.001 | 0.009 |
| PFDA | [-0.009, -0.001] | -0.005 | 0.013 | 0.263 |
| PFOS | [-0.012, -0.001] | -0.006 | 0.019 | 0.263 |
| Interaction |  |  |  |  |
| PFAS | 95% CI | Estimate | p-value | q-value |
| PFOA | [-0.010, 0.001] | 0.0062 | 0.080 | 0.43 |
| PFHxS | [-0.006, 0.006] | -0.0005 | 0.894 | 0.946 |
| PFNA | [0.011, 0.035] | 0.0227 | <0.001 | 0.01 |
| PFDA | [-0.014, -0.0015] | 0.0048 | 0.264 | 0.596 |
| PFOS | [-0.019,-0.005] | 0.0111 | 0.036 | 0.395 |

We then characterized sex-specific relationships via sex-stratified linear regression analysis. We observed consistent directionality across sex strata, where eigengene levels decreased in association with almost all PFAS, but that these effects were stronger amongst female placentas, particularly for PFNA (β = –0.03, p-value = 0.0001) (**Table 5**). We explored whether PFAS concentrations themselves differed between the sexes via t-test comparisons, but none of the 5 PFAS compounds differed significantly between male and female placenta. Together, these results suggested that sexually dimorphic expression in light green was not attributable to PFAS concentration differences and may be due to sex-specific effect modification.

**Table 5:** Sex stratified linear regression analyses examining PFAS as an independent variable and eigengene of the light green module as a dependent variable (adjusted for confounders).

| Male |  |  |  |
| --- | --- | --- | --- |
| PFAS | 95% CI | Estimate | p-value |
| PFOA | [-0.002, 0.004] | 0.001 | 0.423 |
| PFHxS | [-0.003, 0.002] | 0.000 | 0.860 |
| PFNA | [-0.008, -0.0003] | -0.004 | 0.045 |
| PFDA | [-0.006, -0.0001] | -0.003 | 0.029 |
| PFOS | [-0.005, 0.003] | -0.001 | 0.536 |
| Female |  |  |  |
| PFAS | 95% CI | Estimate | p-value |
| PFOA | [-0.012, 0.002] | -0.005 | 0.174 |
| PFHxS | [-0.009, 0.009] | 0.0001 | 0.982 |
| PFNA | [-0.038,-0.013] | -0.026 | <0.001 |
| PFDA | [-0.018,0.001] | -0.008 | 0.092 |
| PFOS | [-0.022, -0.0004] | -0.011 | 0.042 |

### Characterizing the PFAS-associated Module – Light Green

Last, we aimed to characterize the expression patterns, GO terms, and upstream TFs that are important to the only PFAS-associated module, light green. T-test showed that males clearly had a significantly higher eigengene levels compared to females (p = 2.2e-16) and there was a distinct separation in individual gene expression patterns between the sexes in an expression heatmap (**Figure 3**). Interestingly, while clear sex-specific expression patterns were expected and observed for sex-linked genes, the sex differences in expression also extended to the autosomal genes featured in light green. The GO terms associated with light green were largely representative of histone modification processes (**Figure 4**). When using REVIGO to summarize redundant terms affiliated with biological processes, this module was assigned regulation of insulin growth factor receptor signaling pathway (p-value < 0.05). Because TFs can have broad effects on gene activity and may provide insights into co-expression patterns, we identified TFs that are linked to the expression of the genes in the light green module. Placental TF enrichment analysis identified 2 TFs with overrepresented gene-targets in light green (q-values < 0.05) (**Table 6**). *VDR* targeted 5 light green genes (*KDM5C*, *KDM6A*, *UTY*, *EIF1AX*, *EIF2S3*), while *ZFX* targeted a total of 6 genes (*KDM5C*, *KDM6A*, *UTY*, *EIF1AX*, *EIF2S3*, *VDR*).

**Figure 3:**
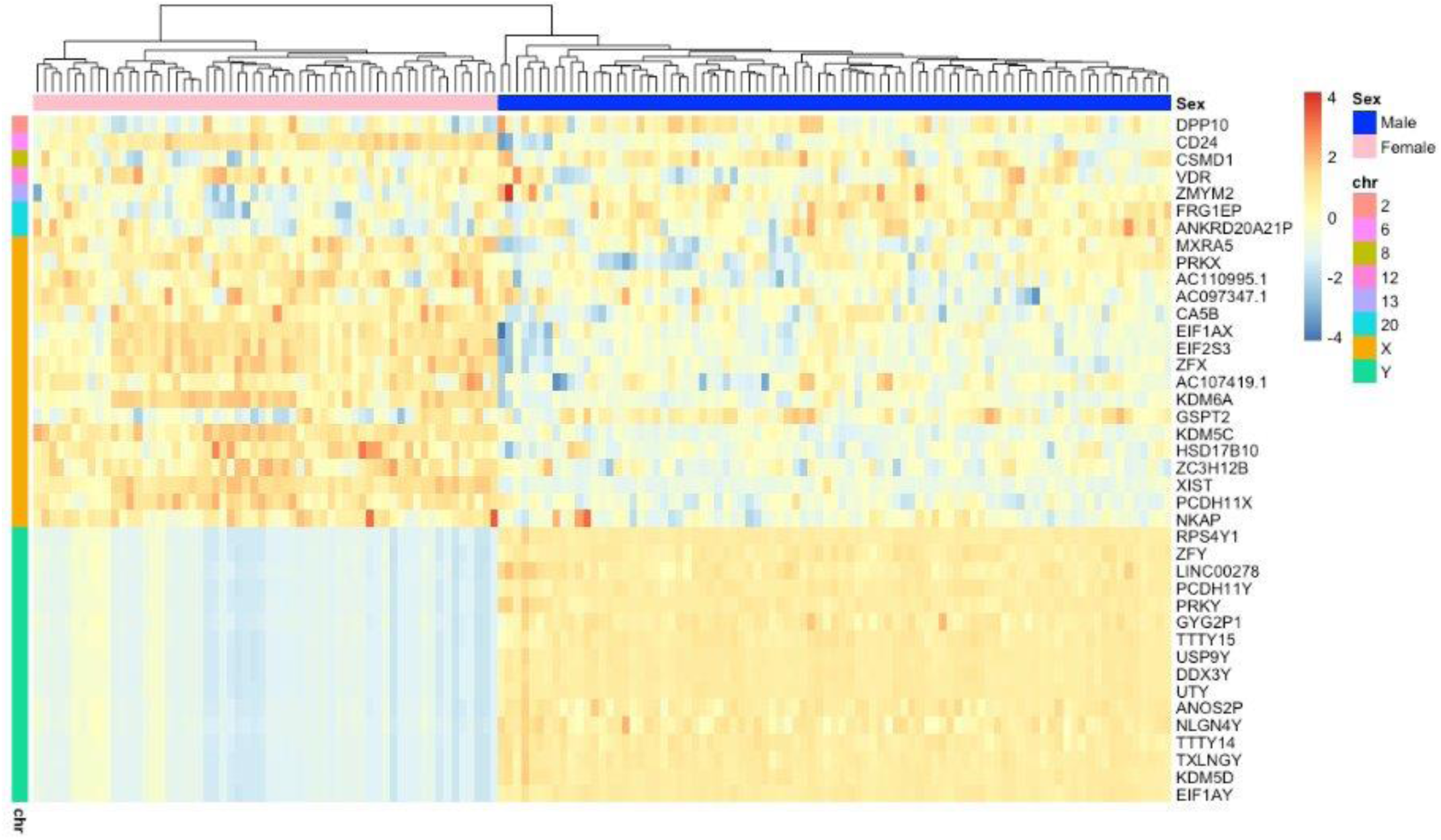
Heat map of expression profiles of light green genes for each participant distinguished by sex and chromosome.

**Figure 4:**
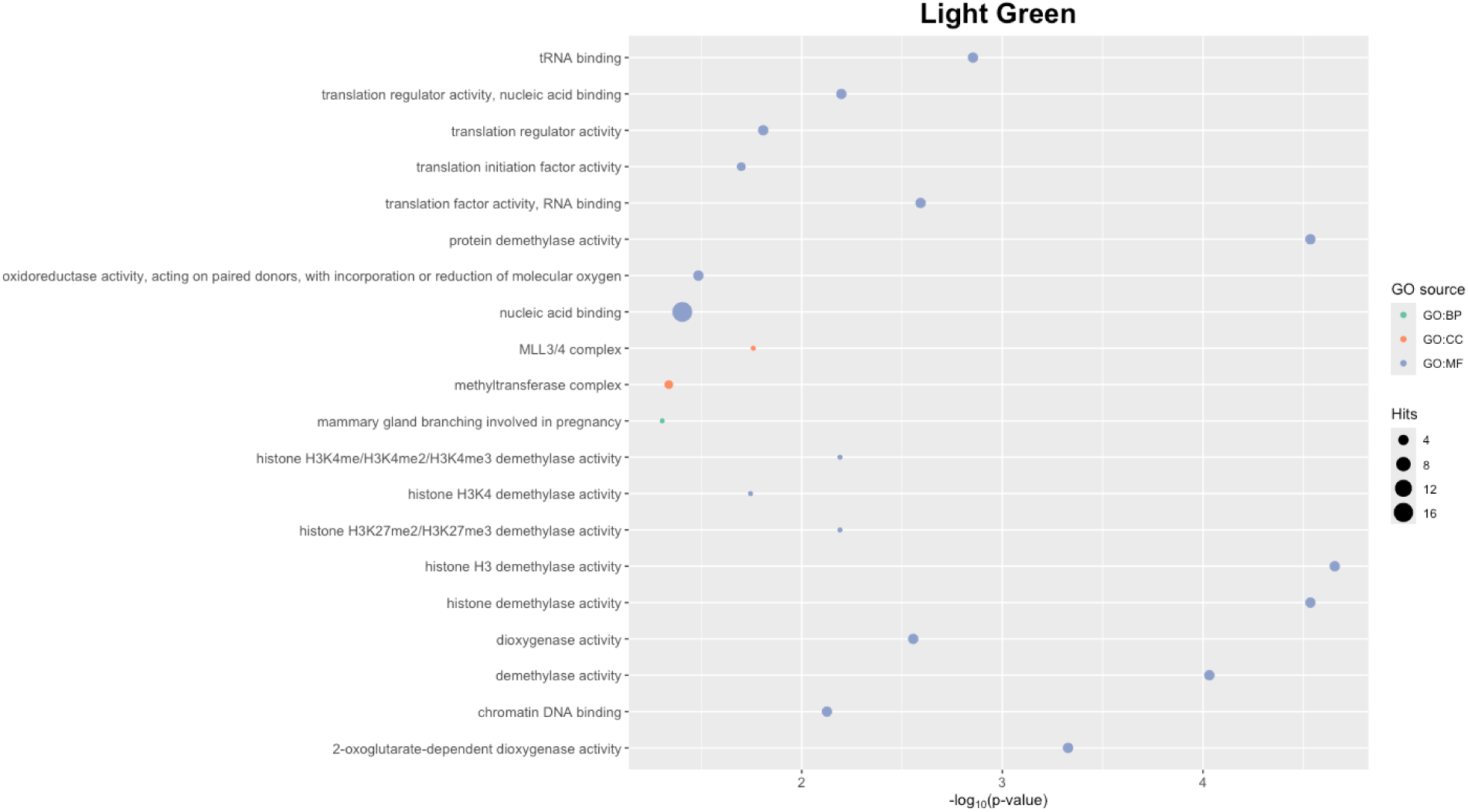
The significant GO terms for light green module are mainly representative of histone activity. The circles indicate the number of genes/hits corresponding to the term and the color relates to the GO source.

**Table 6:** Light green enriched placenta specific transcription factors identified using fisher exact test and adjusted for multiple testing correction.

| TF | p-value | q-value | Light Green Gene Targets |
| --- | --- | --- | --- |
| VDR | 1.76E-07 | 3.52E-07 | *KDM5C, KDM6A, UTY, EIF1AX, EIF2S3* |
| ZFX | 4.57E-06 | 4.57E-06 | *KDM5C, KDM6A, UTY, EIF1AX, EIF2S3, VDR* |

## Discussion

In this study, we applied WGCNA to build a gene network using placental RNA-sequencing data to identify gene expression patterns linked to PFAS exposure. Our analysis revealed a PFAS-related module termed “light green” that was characterized predominately by sex-specific genes and enriched for genes related to histone modification and tissue-specific gene targets of the transcription factors *VDR* and *ZFX*. Prior applications of WGCNA to human placental RNA sequencing data have uncovered modules associated with various birth outcomes and environmental exposures.^42, 43^ While other co-expression studies have identified sex-specific modules, these prior studies did not identify sex-specific associations with exposure and outcomes. ^42–45^ The sex-related modules identified in prior research showed enrichment for biological processes involving DNA conformational changes and post-transcriptional modifications.^42–45^ Our sex-specific light green module was also enriched for some of these same molecular processes. Notably, several terms related to histone modifications were overrepresented within light green, and histone modifications can switch chromatin structure between condensed and relaxed states, influencing DNA conformation.^46^ Furthermore, post transcriptional modifications are often affected by RNA binding, another reoccurring theme in the enriched GO terms for the light green module. Thus, the light green module supports prior analyses that sex differences contribute to tissue specific transcriptional regulation.^31, 47^

Most of the light green genes identified in our investigation are involved in gene expression regulation and have known X-Y homologs.^48^ The X-Y pairs in light green include: *PRKY/PRKX, KDM6A(UTX)/UTY*, *KDM5C/KDM5D*, *EIFAX/EIF1AY*, and *ZFX/ZFY*.^48^ These X homologs are not subject to X-inactivation, leading to higher expression in females. X-inactivation, vital for balancing gene dosage between sexes in mammals, is influenced by factors such as the *XIST* gene, expressed from the inactive X chromosome and present in light green.^49, 50^ Genes active on the ‘silent’ X chromosome are thought to contribute to sex differences and are particularly significant for brain function.^51^

While most studies of prenatal PFAS exposure have not focused on sex-specific effects, several studies have found males and females may be differentially impacted by PFAS. For instance, neurobehavioral disorders can differ in severity by sex and have been associated with PFAS.^52^ Specifically, PFOS and PFOA demonstrate neurotoxic traits, although the connection between PFAS and neurodevelopment remains unclear.^8, 53^ However, there is more consistent evidence linking PFAS exposure to cardiovascular diseases in adulthood. Emerging research suggests that genes on the inactivated X chromosome might also affect the cardiovascular system.^54, 55^ While our study does not link X-Y paralogs and their expression with disease, we identified a statistically significant interaction between PFNA exposure and sex for the light green module eigengene, which indicates that the biological sex of placenta could be key in understanding how PFNA contributes to developmental outcomes. This is especially important considering that sex differences in gene expression apparent in early and term placentas are correlated with sex differences in adult tissues, specifically in the brain.^31, 56^

Although emerging evidence supports that PFAS may have several sex-specific effects, investigations exploring the effects of PFAS exposure on sex-linked gene expression remain limited. To our knowledge, *DDX3Y* is the only light green sex-linked gene with previously reported PFAS associations. *DDX3Y* was upregulated in the livers of male mice fed a high fat diet and exposed to PFNA.^57^ Research into the association between gonosomal genes and PFAS could be limited because of over representation of males in biomedical research studies.^58^ However, many research studies have speculated that PFAS can impact the function of sex chromosomes because PFAS can impair fertility, menstruation, and spermatogenesis.^59–61^ All processes related to sex are ultimately modulated by sex-linked genes; however, there is no research suggesting that PFAS can alter the expression of these genes directly.

While the light green module is composed primarily of X-linked and Y-linked genes, there are also a handful of autosomal genes, most notably the gene for the vitamin D receptor, *VDR. VDR* is the only light green gene that has been previously associated with PFAS exposure. In silico and in vitro assays have shown that PFNA binds to *VDR* and changes the activity of VDR’s target genes.^62, 63^ Additionally, a recent large study of adults showed that increasing levels of PFAS exposure was associated with decreased serum levels of vitamin D, but only among females.^64^ Vitamin D insufficiency has also previously been shown to decrease fetal growth.^65^ In our study, *VDR* expression itself was not associated with any individual PFAS, but *VDR* was within the light green module along with several of its target genes (*KDM5C*, *KDM6A*, *UTY*, *EIF1AX*, *EIF2S3*). Importantly, *VDR* is a transcription factor that mediates the actions of 1,25(OH)_2_D (the biologically active form of vitamin D) including driving transcriptional responses in several tissues, including the placenta.^66^ Our study contributes to the growing body of evidence that the molecular mechanisms involved in vitamin D metabolism are potential targets of PFAS, and that these responses may affect male and female placenta differently.

This investigation reports innovative findings about sex-specific placental transcriptional responses to PFAS, but these findings should be interpreted within the context of some limitations. We identified some autosomal gene expression associations with PFAS, but these did not pass significance thresholds that were corrected for multiple testing, indicating a potential lack of power to detect small effect sizes that was exacerbated by the necessity of sex stratification to study sex-specific effects. The demographic composition of our study sample may limit the generalizability of our results, and since placental samples were collected between 2010-2014, we did not detect high enough concentrations to study the impact of more recently emergent PFAS chemicals. Nonetheless, this cohort is still one of the only to quantify PFAS in placenta with composite birth outcome and molecular omics data. We showcase that even healthy pregnancies can be affected by in utero exposure to PFAS.

The association of the light green module with PFNA suggests that PFNA contributes to sexual dimorphic responses by interacting with sex-linked genes. Light green genes were found to contain X-Y paralog pairs and genes crucial for transcriptional regulation known to influence reproduction, cardiovascular health, and neurodevelopment. The module also was enriched for the TF *VDR,* a gene represented within light green that has also been previously associated with PFAS exposure. Overall, our study provides novel findings that higher levels of placental PFNA were associated with downregulation of some placental genes, but only among female placentas. These results were consistent with our sex-stratified differential expression analysis where only females exhibited a response. Some of the light green genes are targets of *VDR*, implicating changes to vitamin D metabolism and downstream pathways, which may be related to the genes within the module that are broadly involved in insulin growth factor receptor signaling and histone modifications. Further research is needed to understand whether these gene expression responses to PFAS are related to placental function, pregnancy outcomes, and children’s health outcomes.

## Data Availability

All data produced in the present study are available upon reasonable request to the authors.

