## Supplemental Figures 1-3 for "Sex-Dependent Relationships Between PFAS and Placental Transcriptomics Identified by Weighted Gene Co-Expression Analysis"

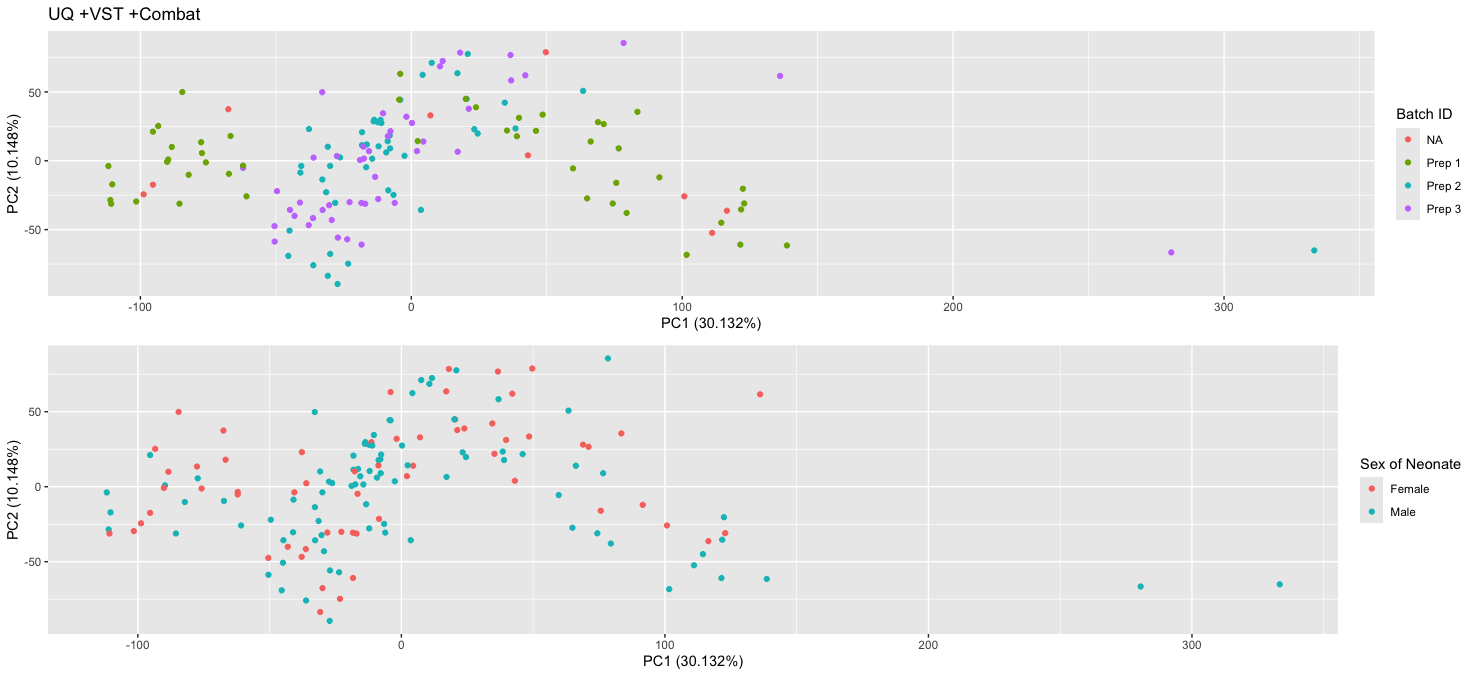

**Supplemental Figure 1:** PCA plots annotated by batch or sex. Some batch IDs were labeled as NA if not reported. No clear subgroups were observed in our data. Sex of neonate is our primary variable of interest. We conducted a Wilcoxon test to determine if PC1 differed by sex and reported no significance (p-value > 0.05).

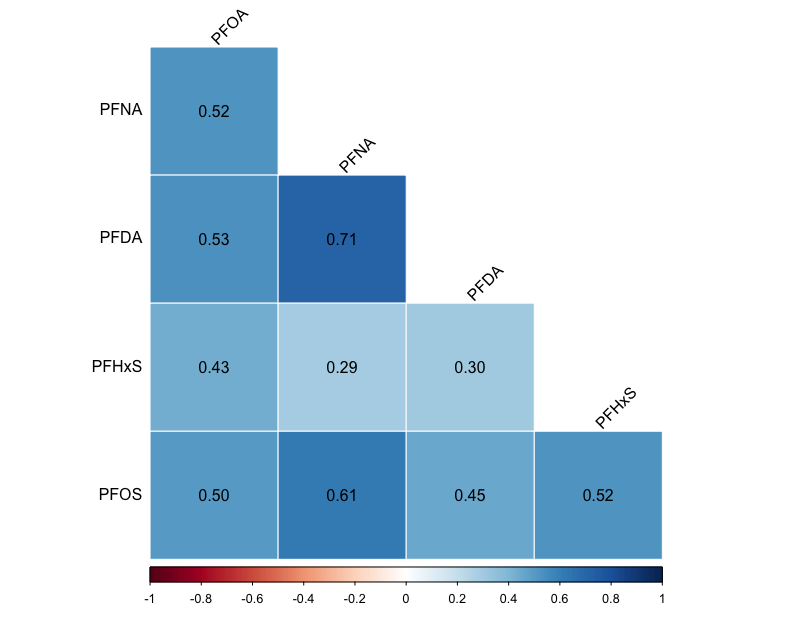

**Supplemental Figure 2:**  A spearman correlation heatmap of the most detected PFAS among the 147 Glowing participants (p-value < 0.05).

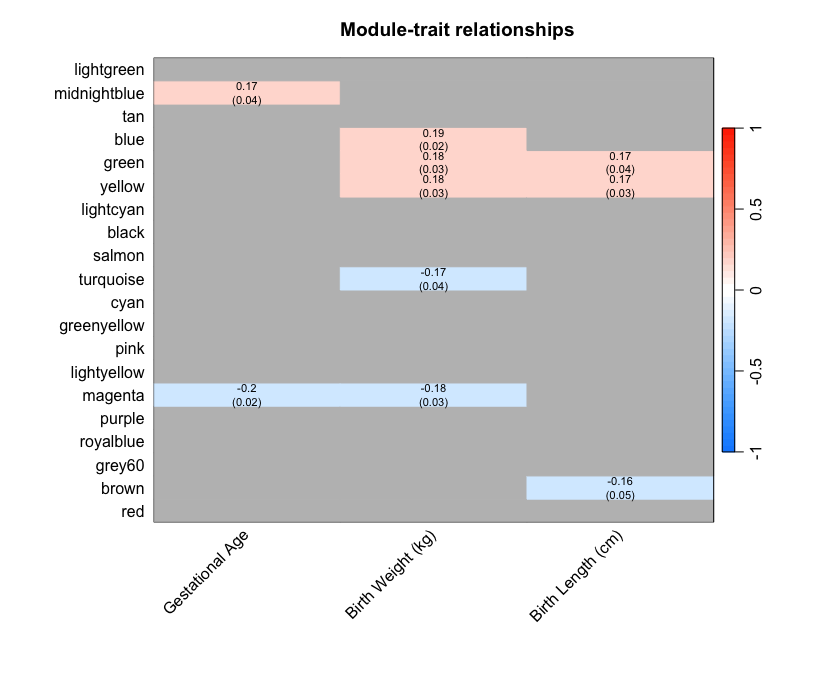

**Supplemental Figure 3:** Pearson correlation analysis of modules with a significant association to a birth outcome.
